# The ocular tissue-specificity of differentially expressed age-related macular degeneration associated genes

**DOI:** 10.1101/2020.12.28.20248962

**Authors:** Dylan Duchen, Terri Beaty

## Abstract

**Purpose:** Age-related macular degeneration (AMD) is a leading cause of blindness in the developed world. One of the most genetically well-characterized degenerative diseases, genome-wide association studies (GWAS) have identified 52 independent common or rare AMD risk associated variants. While transcriptome-wide association analyses (TWAS) and expression quantitative trait loci (eQTL) efforts have characterized the effects of these AMD-associated genes on mRNA expression in retinal tissue, we aimed to characterize the AMD-associated transcriptional profiles of functionally distinct ocular tissues including the macular and extramacular regions of the retina and the retinal-pigment epithelium (RPE)/choroid.

**Methods:** Using publicly available microarray data (NCBI GEO accession: GSE29801) comprised of retinal and RPE/choroidal tissue samples from 142 AMD patients and 151 healthy individuals (118 retina and 175 RPE/Choroid samples), tissue-specific differential gene expression analyses were conducted. Transcriptome analyses were focused on 878 genes surrounding known AMD-associated loci.

**Results:** Many genes which contain clinically significant or causal variants identified via GWAS or TWAS/eQTL studies were significantly differentially expressed and display transcriptional heterogeneity across different subtypes of ocular tissue and retinal geography in AMD-associated tissues.

**Conclusion:** These findings demonstrate the importance of spatial heterogeneity and tissue specificity in the mRNA expression of known AMD-associated genes. Genes known to harbor rare or causal AMD-associated variants are differentially expressed in functionally distinct ocular tissues of AMD patients, suggesting they might contribute to disease regardless of mutation status.

## Introduction

Age-related macular degeneration (AMD) is one of leading causes of blindness in the developed world and is a growing problem in developing countries where average life expectancies are increasing.^1^ Based on several population-based cohorts, the estimated prevalence of AMD is 0.2% in people aged 55–64 years and rises to 13% in people older than 85.^2^ While a range of conditions may result in macular degeneration, AMD reflects the progressive accumulation of drusen, deposits of proteins, lipids, and trace elements, which aggregate within and beneath the retina and the retinal pigment epithelium (RPE). AMD presents in three distinctive categories depending on the severity of vision loss; early AMD, intermediate AMD, and advanced AMD. Advanced AMD results from regions of geographic atrophy, the loss of focal areas of the RPE and its supporting blood vessels, or neovascularization. Neovascular AMD has an acute onset and can lead to detachment of RPE tissue or the retina, resulting in severe central vision loss.^3^ Additional classifications exist, including ‘wet’ (choroidal neovascularization with angiogenesis) and the more common ‘dry’ AMD (geographic atrophy without angiogenesis).

European-ancestry, in addition to age and tobacco use,^2,3^ is a major risk factor for developing AMD. These ancestry-related differences persist despite those of European-ancestry having the same prevalence of drusen deposits as other ethnicities.^4–7^ A role for genetics in AMD development was initially supported by familial aggregation studies,^8^ with the *complement factor H* (*CFH*) gene being identified as AMD-associated in one of the earliest genome-wide association studies (GWAS).^9^ Subsequent GWAS have resulted in AMD being one of the most genetically well-characterized degenerative retinal diseases.^10–16^ The most recent GWAS involved a cohort of 16,144 AMD cases and 17,832 controls, testing over 12 million variants for association with various AMD subtypes.^17^ Bringing the total number of independent variants associated with risk of AMD to 52 (involving 34 different genetic regions), this GWAS identified three genes (*CFH, CFI*, and *TIMP3*) as harboring rare (MAF<0.1%) causal variants and a fourth gene (*SLC16A8*) harboring a potentially causal splice variant and estimated the additive genetic effects of these markers combined accounts for 46.7% of the variability in risk of AMD among those of European ancestry.^17^

While rare causal variants can hint at underlying biological mechanisms of AMD and may completely explain the etiology of AMD for a few individuals, the extent to which rare causal mutation-associated genes affect AMD risk among individuals without these mutations remain unclear. Determining whether these genes are also differentially expressed in AMD compared to normal eye tissue could suggest additional mechanisms by which these genetic factors contribute to the pathogenesis of AMD. While Fritsche, et al. surveyed the mRNA expression for the 368 genes closest to the recognized 34 regions of interest in healthy retinal or choroidal tissues via RNAseq,^17^ AMD-derived tissues have been shown to have significantly different transcriptome profiles compared to normal ocular tissue.^18–23^ Recently, a transcriptome-wide association analysis (TWAS) and expression quantitative trait loci (eQTL) study comparing diseased to healthy retinal tissues identified additional causal variants across these 34 GWAS-identified risk loci and characterized the effects of AMD-associated variants on gene expression within the retina.^24^

As gene expression is spatially heterogenous and differs across functionally distinct tissues and regions within the eye, these findings may ignore tissue-specific expression differences relevant to AMD pathogenesis. For example, neither the combined GWAS study of Fritsche et al. or Ratnapriya et al.’s TWAS/eQTL study included RPE/choroidal-specific AMD tissues or attempted to distinguish between macular and extramacular regions of the retina (Figure 1).^25^ Thus, it remains unclear whether the genes identified by these large genomic studies are differentially expressed in AMD patients within distinct geographical regions or certain tissue subtypes. Leveraging publicly available data,^23^ we aimed to determine whether these GWAS-associated genes are aberrantly expressed in AMD compared to healthy tissues within retinal and RPE/choroidal tissues. Identifying the distinct tissues and regions in which AMD-associated genetic factors are aberrantly expressed will improve our understanding of AMD pathogenesis and potentially aid drug development efforts by revealing novel mechanisms by which genetic factors contribute to disease.

**Figure 1:**
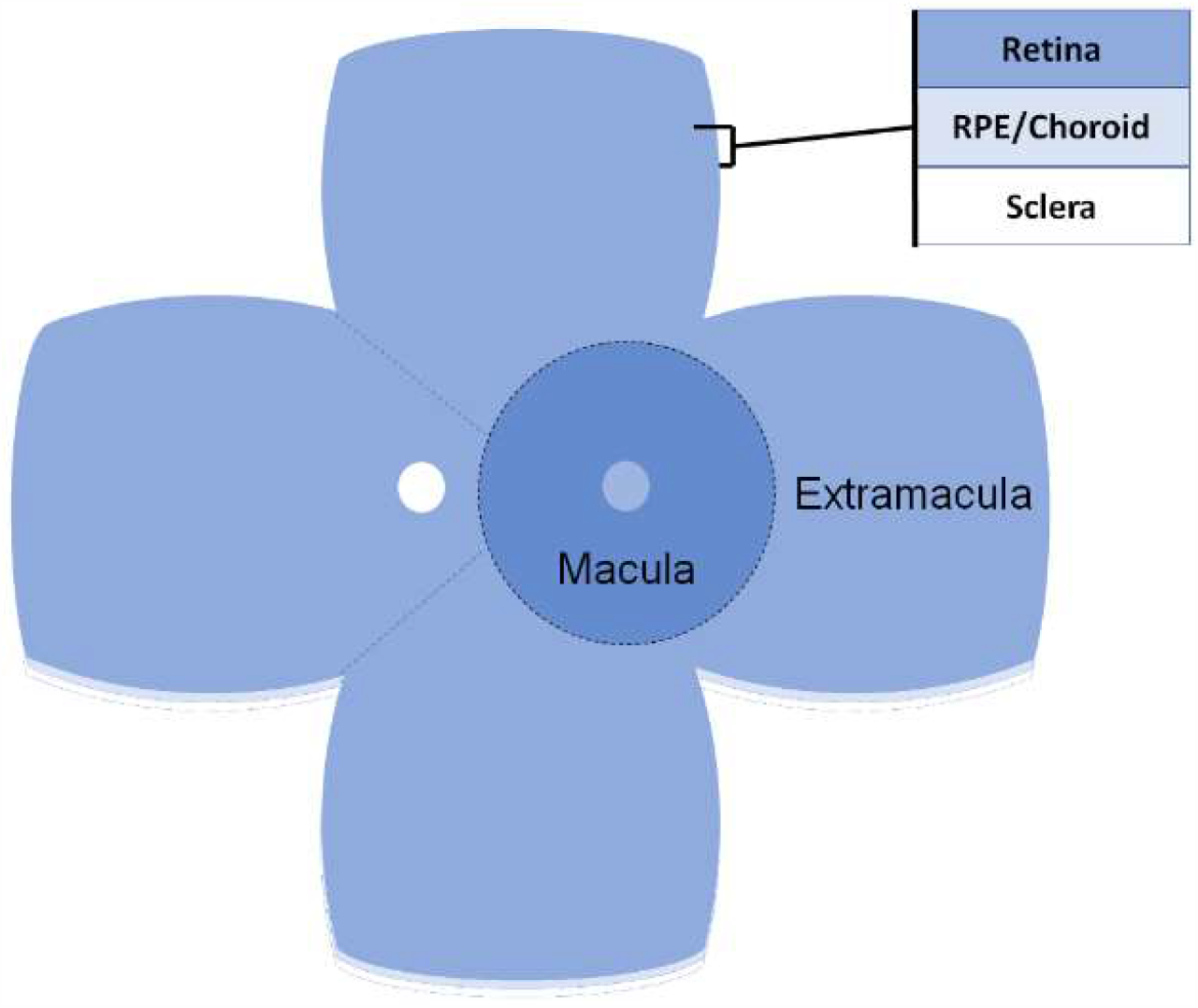
Ocular tissue layers and geographical regions relevant for AMD-associated transcriptome analysis. Figure adapted from Morgan et al.^25^

## Materials and methods

Publicly available transcriptome data was obtained through NCBI’s Gene Expression Omnibus (GEO), accession GSE29801, using the GEOquery (v2.48.0) package in R. Tissue acquisition, RNA purification, and microarray hybridization methods have been described previously.^23,26^ The two channel Agilent Whole Genome 4×44 K *in situ* oligonucleotide array platform (G4112F, Agilent Technologies, Inc., Santa Clara, CA, USA) was used to assess global transcriptome mRNA expression utilizing a pooled reference panel consisting of equal parts RPE-choroid and retinal RNA. A dye-swap design was implemented where samples from the pooled reference and tissues of interest were labeled with alternating dye-labels to minimize the chance of systemic dye effects. During the preprocessing stage, experimental and pooled reference samples were assigned to the appropriate channels as if no dye-swap were conducted, benefiting from the dye-swap design without requiring any downstream dye-related normalization steps. Samples were background corrected by subtracting the background intensities from the foreground intensities, as previously described.^23^ Within-array normalization was conducted via loess normalization followed by quantile normalization across arrays, according to the limma (v3.36.3) differential gene expression-focused software package.^27^

Negative and positive control probes as well as the lowest expression probes were filtered out by computing the 95^th^ percentile of the negative control probes for each array, and then retaining probes which were at least 10% brighter than these values in at least four arrays. Duplicate probes (N= 253) within each array were collapsed by their mean, resulting in a total of 33,237 unique probes. Probe-to-gene annotation was accomplished using the Agilent “Human Genome, Whole” annotation package hgug4112a.db (v3.2.3) accessed via the Bioconductor suite of R packages.^28^ As our intended level of inference is the gene level, the probe with the highest inter-quartile range across all samples was selected to represent genes containing multiple mapped probes, resulting in 16,716 unique genes available for analysis. Differential gene expression analysis was conducted by first stratifying by tissue-source (macular region, extramacular region, retina, RPE/choroid, macular retina, macular RPE/choroid, extramacular retina, and extramacular RPE/choroid) and then fitting linear models for the mRNA expression of each gene with the empirical Bayes method according to the limma differential gene expression guidelines.^27,29^

Written consent was obtained from all participants or their surviving relatives under protocols which adhered to the guidelines described by the Declaration of Helsinki, as previously described.^23^ This previous study was approved by the St. Louis University, the University of Iowa, the University of Utah, and the University of California, Santa Barbara institutional review boards.^23^

## Results

The large GWAS conducted by Fritsche et al. identified 34 loci achieving genome-wide significance, corresponding to regions containing 878 different genes, defined by linkage-disequilibrium (LD) (r^2^≥ 0.5, ± 500 kb).^17^ Of these 878 genes, 650 were annotated and expressed (74%) in our analysis of RNA expression. Using a narrower LD defined region surrounding the 52 most significantly associated markers (r^2^≥ 0.5, ± 100 kb), 368 unique genes were located within these regions and used by Fritsche et al., for subsequent analyses. Of these 368 genes, 274 (74.5%) were annotated and found to be expressed in our analysis of RNA expression in 8 different retinal tissues.

After stratifying by both tissue origin and region, 289 genes were differentially expressed (FDR< 20%) within the macula of the retina, while in RPE/choroid tissues more genes in the extramacular region were differentially expressed (a total of 889 genes) compared to the macula (only 86 genes). Combining macular and extramacular retinal samples, differential gene expression analyses identified 2,491 differentially expressed genes (*p*< 0.05) in retinal tissues (N= 118), 590 with FDR< 10%. Of these genes, 123 were also identified in the large GWAS study, 58 of these genes located within the narrow windows of LD surrounding 34 recognized risk loci. Twenty-nine of the GWAS-identified genes included within the wider LD cutoffs were significantly differentially expressed between AMD and healthy tissues at a FDR< 10%, of which the following 17 genes fell within the narrowly LD-defined subset of 368 different genes: *C4B, CFB, HLA-B, HLA-DQA2, TMEM116, HLA-C, APOE, C3, HLA-DRA, HTRA1, CLIC1, MMP19, ABCA1, PLTP, TRPM3, TNXB*, and *HSPH1* (Table 1). The three genes (*HLA-DQB2, CYP21A2*, and *GALR3*) were determined to not be expressed in healthy retinal tissue according to the RNAseq analysis conducted by Fritsche et al. yet were significantly differentially expressed in AMD vs. healthy retina tissue in this analysis.^17^ *CFI* and *TIMP3*, two of the three genes identified as potentially harboring causal rare variants (i.e. those with MAF< 0.1%), were also differentially expressed in AMD-diagnosed retinal tissue (i.e. showing higher expression in AMD compared to healthy retina with FDRs of 12.7% and 17.5% respectively).

**Table 1:** Differentially Expressed Genes Stratified by Tissue and Geographic Region. Shaded rows indicate retina-specific tissue findings.

|  |  | *Differentially Expressed Genes ($p < 0.05$)* |  |  | *Differentially Expressed Genes ($FDR < 10\%$)* |  |  |  |  |
| --- | --- | --- | --- | --- | --- | --- | --- | --- | --- |
| Tissue | Samples (N) | Genes (N) | Overlapping GWAS-Identified Genes (Wide Margin, N=878) | Overlapping GWAS-Identified Genes (Narrow Margin N=368) | Genes (N) | Overlapping GWAS-Identified Genes (Wide, N=878) | Overlapping GWAS-Identified Genes (Narrow, N=368) | Overlapping GWAS-Identified Genes (DE with $FDR < 10\%$) | Overlapping TWAS-Identified Genes (DE with $FDR < 20\%$) |
| Retina | 118 | 2491 | 123 | 58 | 590 | 29 | 17 | *C4B, CFB, HLA-B, HLA-DQA2, TMEM116, HLA-C, APOE, C3, HLA-DRA, HTRA1, CLIC1, MMP19, ABCA1, PLTP, TRPM3, TNXB, HSPH1* | *CFB, APOE, PLTP, HLA-DQB1, C2, CFI, COL8A1, TMEM199, CD63, HERPUD1, NIT2* |
| RPE/Choroid | 175 | 2452 | 104 | 52 | 443 | 20 | 10 | *SLC25A10, COL8A1, HCP5, SRPK2, MRPL12, HLA-DRB1, APOC1, GDF11, PILRB, VEGFA* | *COL8A1, SRPK2, PILRB, HERPUD1, PLA2G12A, SLC16A8* |
| Macular Retina | 60 | 1766 | 70 | 36 | 70 | 4 | 2 | *C4B, C3* | *APOE, PLTP* |
| Macular RPE/Choroid | 91 | 1786 | 75 | 35 | 11 | 4 | 1 | *PBX2* |  |
| Extramacular RPE/Choroid | 84 | 2349 | 108 | 48 | 302 | 14 | 7 | *DDAH2, BLOC1S1, PILRB, COL8A1, ARHGAP21, HCP5, APOM* | *BLOC1S1, PILRB, COL8A1, TOMM40, NIT2* |

|  |  |  |  |  |  |  |  |  |  |
| --- | --- | --- | --- | --- | --- | --- | --- | --- | --- |
| Extramacular Retina | 58 | 1398 | 70 | 36 | 0 | 0 | 0 |  |  |
| Macula (retina + RPE/Choroid) | 151 | 1367 | 58 | 25 | 5 | 0 | 0 |  |  |
| Extramacular (retina + RPE/Choroid) | 142 | 1212 | 63 | 31 | 2 | 0 | 0 |  | *PILRB* |

In RPE/choroid tissue (N= 175), a total of 2,452 genes were differentially expressed (at *p*< 0.05), and 443 yielded a FDR< 10%. Among these genes, 104 had been previously identified within the wider LD regions surrounding the 34 most significant genes by Fritsche et al. while 52 were within the narrow LD regions. Ten of these genes were significantly differentially expressed between AMD and healthy tissues (defined as FDR< 10%): *SLC25A10, COL8A1, HCP5, SRPK2, MRPL12, HLA-DRB1, APOC1, GDF11, PILRB*, and *VEGFA* (Table 1). Comparing mRNA expression results generated here to the RNAseq results from healthy RPE/choroid tissues conducted by Fritsche et. al., seven genes (*UGT3A2, TNXB, BTNL2, HLA-DQA2, HLA-DOB, CKM, TIMP3*) were discordant in that they showed differential expression in AMD vs. healthy RPE/choroid (*p*< 0.05) while they were not expressed in the RNAseq analysis of healthy retina/choroid tissues. As noted above, *TIMP3* (*p*= 0.039, FDR= 31.2%) had been previously identified as potentially containing rare causal variants (MAF< 0.1%).

Genes included within the narrow-LD regions surrounding the recognized GWAS risk loci identified as significantly differentially expressed (at FDR< 10%) in extramacular RPE/choroid tissue (N= 84) included: *DDAH2, BLOC1S1, PILRB, COL8A1, ARHGAP21, HCP5*, and *APOM*, all of which were reported to be expressed by Fritsche et al.’s RNAseq analysis. Additional findings for macular-specific retina, macular-specific RPE/choroid, extramacular-retina, macular retina and RPE/choroid combined, and extramacular retina and RPE/choroid combined are shown in Table 1. Using a relatively loose FDR threshold of 20%, the narrow-LD region defined genes *COL8A1, HCP5, APOM, HSPA1A, HLA-DQA2, VEGFA*, and *HERPUD1* showed differential expression in both retina and RPE/choroid tissue.

Attempts at characterizing mRNA expression of AMD-associated genes is not a novel concept.^25^ Comparing our results to a recently conducted TWAS and expression quantitative trait loci (eQTL) analysis in bulk retina tissue,^24^ 10 significantly differentially expressed genes (FDR< 20%), including *PLTP, HLA-DQB1, CFI, TMEM199, HERPUD1, NIT2, PILRB, PLA2G12A, BLOC1S1*, and *TOMM40* were previously identified as either a target gene of causal variants or was found to contain TWAS/eQTL-associated variants. Genes identified in Ratnapriya et al.’s eQTL/TWAS analysis which were differentially expressed (FDR< 20%) solely within the retina include *CFB, APOE, PLTP, HLA-DQB1, TMEM199, CFI, C2*, and *CD63*. Differentially expressed eQTL/TWAS-identified genes specific to RPE/choroid include *BLOC1S1, SRPK2, PLA2G12A, SLC16A8, TOMM40*, and potentially *PILRB* (all of these genes were differentially expressed within RPE/choroid tissues, extramacular RPE/choroid tissues, as well as extramacular tissues of retina and RPE/choroid combined). Genes differentially expressed across both retina and RPE/choroid were emphasized in the eQTL/TWAS analysis and included *COL8A1, HERPUD1*, and *NIT2. B3GLCT*, one of two genes strongly suggested as being causal for AMD pathogenesis in the TWAS/eQTL study, was not included in these results because *B3GLCT*-specific probes were not included in the Agilent annotation package. Individual probe-level analysis of probes mapping to *B3GLCT* showed a marginal difference in expression in retinal AMD tissues (*p*= 0.03, FDR= 27.5%). However, the gene immediately upstream of *B3GLCT, HSPH1*, was included within the narrow-LD regions surrounding the top GWAS loci and was significantly differentially expressed in AMD-associated retina tissue (FDR< 10%).

## Discussion

The retina includes 10 distinct layers serving varying biological functions and is composed of specialized cells and substructures for photoreception and transmitting signals through the optic nerve. The RPE, a single layer of epithelial cells supporting the neural retina with nutrients, is the basal layer of the retina and is most distal from the vitreous filled interior of the eye. In addition to these layers being comprised of multiple, disparate cell types, the distribution of certain cells is heterogenous across the retina. For example, rod and cone cells, photoreceptors responsible for scotopic (low light, low acuity, and non-color vision) and photopic (bright light, high acuity, and color vision) respectively, vary in density depending on their proximity to the center of the macula. When the composition and the function of a tissue varies spatially, so too does RNA transcription, transcriptional regulation, and the translation of genetic material. The two main tissue types, the retina and RPE/choroid, as well as their respective regions (the macular and extramacular regions) are important for development and progression of AMD. Determining whether the contribution of recognized genes associated with risk to AMD is spatially or retinal tissue-type specific should improve our understanding of the biological mechanisms underlying AMD development.

Genes differentially expressed between retina and RPE/choroid tissues, (e.g., *HERPUD1, COL8A1, HCP5, APOM, HSPA1A, HLA-DQA2*, and *VEGFA*) may contribute to AMD pathogenesis by uniformly affecting several tissue subtypes. However, when gene expression spatially differs between retinal and RPE/choroid tissues (as it does for many genes associated with AMD) a potential interaction between geographic region and tissue layer may exist. Because the macular region of the retina and extramacular region of the choroid contain the most differentially expressed genes (in their respective tissue types), it suggests a region/tissue specificity of the genetic architecture and pathogenesis of AMD.

These tissue and region-specific findings allow discrimination between retina or RPE/choroid-specific signals, potentially providing evidence supporting the hypothesis that these signals may be more relevant in the macular or extramacular regions. *BLOC1S1*, for example, was a TWAS-associated variant containing gene and the gene targeted by the lead causal variant located within the *RDH5/CD63* region. In this analysis, *BLOC1S1* was differentially expressed only within the extramacular RPE/choroid (Figure 2). Encoding a component of the BLOC1 protein complex required for the biogenesis and function of the endosomal-lysosomal system, *BLOC1S1* was previously hypothesized to play a role in synaptic function.^24^ *BLOC1S1*’s putative function as a transporter between endosomes and lysosomes, and the observation that a deficiency in certain subunits of the BLOC1-protein complex can result in accumulation of surface proteins could suggest a role in protein accumulation beneath the extramacular RPE/choroid.^30,31^ Indeed, extramacular drusen are associated with AMD development.^32^ The effect of overexpression of *BLOC1S1* on BLOC1 activity, as well as this putative functional relationship warrant further study. *PILRB*, a gene identified in both the GWAS and TWAS/eQTL studies represents another gene differentially expressed within the RPE/choroid but not the retina, potentially indicating some tissue-specific role for the *PILRB* gene in AMD pathogenesis. We also observed two GWAS identified risk genes for AMD harboring rare non-synonymous variants, *TIMP3* and *CFI*, as differentially expressed in retinal tissues critical to AMD. *TIMP3* was marginally significantly differentially expressed in retina (FDR= 17.5%) and extramacular choroid (FDR= 15.6%) tissues, while *CFI* was significantly differentially expressed in retina (FDR= 12.7%). The characterization of genes containing rare variants (like *TIMP3* and *CFI*) as significantly differentially expressed suggests some expanded functional role for these genes in development and pathogenesis of AMD independent of their mutation status. *CFH*, the third rare-variant containing gene highlighted by Fritsche et al., was not differentially expressed in AMD compared to healthy tissue in either retina or the RPE/choroid, potentially indicating whatever its biological mechanism, *CFH*-associated pathogenicity may not be mediated through altered RNA expression.

**Figure 2:**
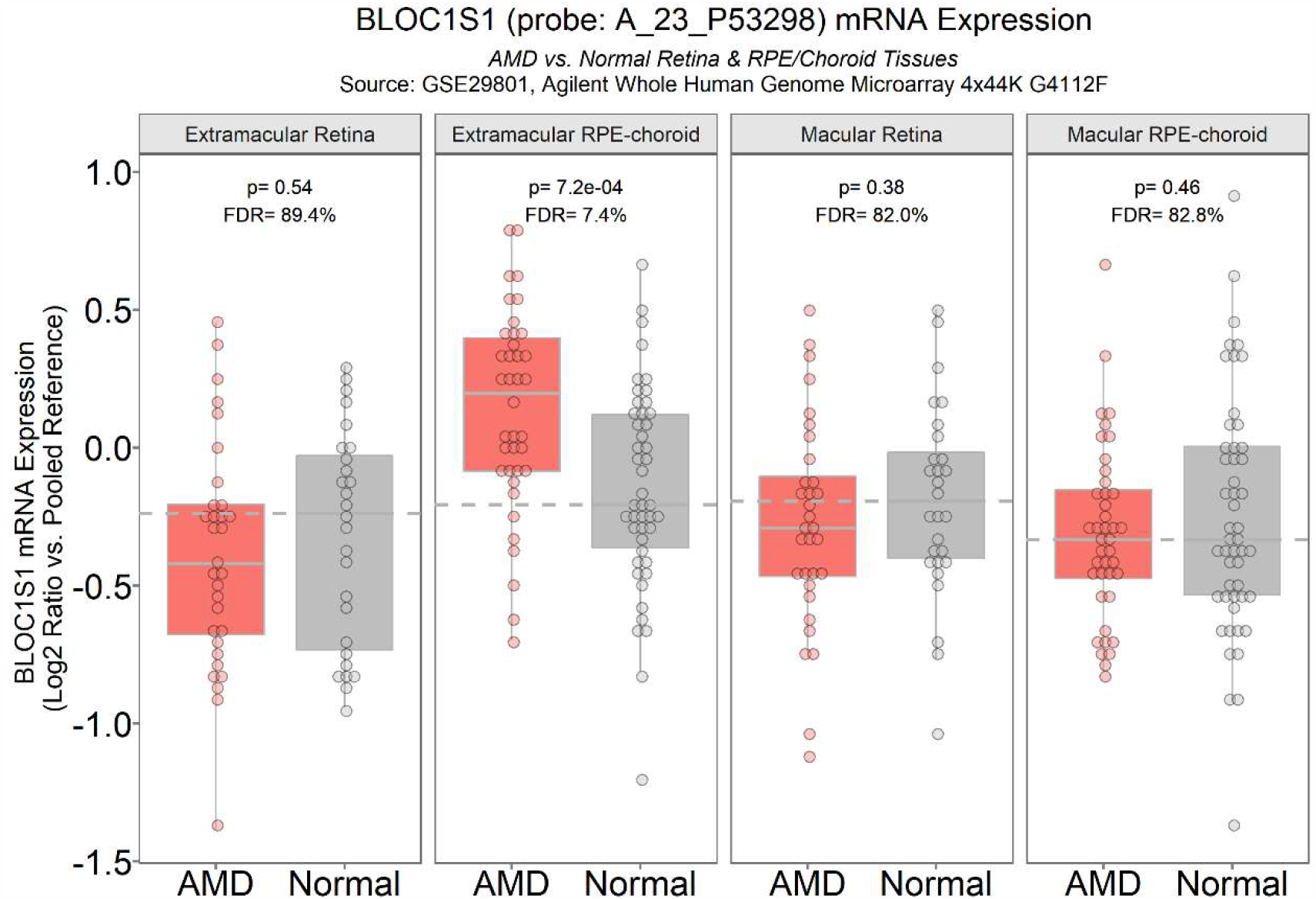
*BLOC1S1* mRNA expression across ocular tissue types. Dotted horizontal lines indicate the median log2-transformed intensity ratio comparing the relevant tissue/region-specific normal samples to the pooled reference.

Information related to genetic ancestry of the donors was unavailable, potentially biasing the results and the generalizability of our findings as transcriptional regulation and mRNA expression can differ significantly depending on ancestry.^34,35^ While we can conclude these donor samples were not included in Fritsche et al.’s GWAS cohort, we cannot guarantee that our donor population is genetically similar to the source populations included in either Fritsche et al.’s GWAS or Ratnapriya et al.’s TWAS. Despite this limitation, the significant amount of overlap between results of this analysis and Fritsche et al. and Ratnapriya et al.’s findings (see Results) suggests the donor samples utilized within this analysis were not genetically distinct from those included in these previous GWAS or TWAS studies, a possibility as a subset of donor samples from Fritsche et al. were also sourced from the University of Iowa and Oregon Lions.^10,17,23,36^

This research focuses on genes previously found to be associated with AMD in large GWAS and TWAS/eQTL studies. These previous studies do not provide the geographic resolution to establish whether differential expression of these genes is specific to unique regions in the eye or distinct tissue layers. These findings highlight both the spatial and tissue-specific contribution of known risk genes for AMD, adding depth to our understanding of the biological importance of these genetic risk factors for AMD. Additional functional analyses are still needed to determine whether these identified regions and/or tissue-specific expression of these genes are truly biologically meaningful or directly pathogenic.

Furthermore, while the prevalence of AMD and subsequently the genetic risk for AMD may be highest in populations of European descent, AMD is certainly not exclusive to this population.^33^ While the effects of genetic variation on RNA expression may be ancestry specific and suffer from the same generalizability issues seen with GWAS studies.^34,35^ Large GWAS and tissue layer-specific eQTL-focused TWAS studies with diverse cohorts are still needed to confirm whether these results hold for individuals of non-European ancestry.

## Data Availability

All data used to support the findings of this study are publicly available in NCBIs Gene Expression Omnibus (GEO), accession GSE29801, at https://www.ncbi.nlm.nih.gov/geo/query/acc.cgi?acc=GSE29801

## Acknowledgements

This work was supported by the National Eye Institute’s Predoctoral Eye & Vision Genomics Training Grant, T32-EY022303. The authors would like to thank the volunteers and patients who participated in the multiple studies from which publicly available microarray and any clinically associated metadata was derived.

## Data availability

All data used to support the findings of this study are publicly available in NCBI’s Gene Expression Omnibus (GEO), accession GSE29801, at https://www.ncbi.nlm.nih.gov/geo/query/acc.cgi?acc=GSE29801.

## Supplemental data

Tissue-Specific Differential Gene Expression (*p*< 0.05) Results

